# Presence of glyphosate in urine due to environmental exposure among populations of agro-industrial areas in Buenos Aires, Argentina

**DOI:** 10.1101/2024.04.02.24305133

**Authors:** Silvana Figar, Analia Ferloni, Amparo Saravi, Adriana R. Dawidowski, Valeria I. Aliperti, Ignacio Bressán, Florencia De Florio, Jimena Vicens, Nahuel Braguinsky Golde, Natalia K. Garcia, Glenda Pazur, Guillermo E. Hough, Adrián C. Gadano

## Abstract

**Introduction:** An increasing number of rural communities express perception of health damage from glyphosate and other agrochemicals. We measure the presence of glyphosate in the human body, in order to create, together with the local community, a systemic model that highlights modifiable causal socio-environmental conditions.

**Materials and methods:** Participatory Action Research. Measurement of environmental factors and self-reported oncological disease were obtained in a three-stage probabilistic sampling (blocks, houses, family) of people living in French city - 9 de Julio-Buenos Aires. Glyphosate in urine was analyzed by liquid chromatography coupled to tandem mass spectrometry. The exposure pathway was obtained by interviewing positive cases. A conceptual systemic model was designed.

**Results:** of the total 46 blocks of French, 23 were included with systematic sampling and from the 76 houses selected (50%) one person was included in the study. Oncological disease was reported in 21.8% of the households. 13% of the population (95% CI 6.5-23) presented quantifiable glyphosate in urine in June 2023. Occupational exposure was ruled out in all cases. The main self-reported sources were: unloading agrochemicals in the nearby warehouse, the grain storage complex, pesticide drift and self-propelled sprayers that pass by on the street, variables belonging to economic and cultural conditions. A network of actors emerged who, gathered on a website, propose actions to the mayor’s office.

**Discussion:** This study has high external validity for public health decision makers regarding the determinants. It is necessary to notify the Argentine Integrated Health System, both suspected exposure and possibly related health events, and to design how to refer human samples to highly complex laboratories to measure pesticides.

**Conclusion:** the presence of glyphosate in urine was due to environmental exposure; It expresses a path of passive, involuntary and chronic absorption of environmental pollutants and is due to French’s agricultural activity with dominance of market forces in the system, poorly antagonized by care forces.

## Introduction

Since glyphosate was discovered as a herbicide in 1970, new mechanisms of action have been described, the consequences of which for humans are hardly attributed solely to glyphosate following the classic risk model.^1-2^. This is because these consequences can manifest themselves in the absence of the exposure factor long after having acted during embryogenesis or the window period.^2–4^.

In March 2015, the International Agency for Research on Cancer announced that glyphosate should be considered as probably carcinogenic^5^; classification which has been adopted by Argentina’s National Cancer Institute. Although new evidence has recently been published on the negative health effects of glyphosate on animal and human health^6-7-8^, this classification is still in doubt, as can be seen in the recent EFSA report^9^. Thus, in the face of these controversies, the joint approach of regulatory agencies, civil society and scientists is essential to make informed decisions in the context of uncertainty.

In the recent SPRINT study, which covered 17 countries, the highest measurements of these pesticides, in the environment and in human biological matrices, were observed in Argentina^10^. There is cumulative toxicity in animals due to the daily pesticide load coming from feed, with documented synergic interaction in sub-threshold doses of different pesticides being present together^11^. These studies, and the evidence that glyphosate is an endocrine disruptor^4,12^, show that little is known on the long-term impact of multiple pesticides co-exposure at reproductive level and during humans’ development. The highest consensus level is on evidence of Non-Hodgkin’s lymphoma in humans^4,12,13^.

Another general effect of pesticides, which appears with increasing frequency, is their contribution to multi-resistant bacteria^14^. Recently, the effect of glyphosate on the microbiome was demonstrated by its selection of pathogenic bacteria that survive the minimum inhibitory concentration of several antibiotics^15^. Glyphosate interferes with the production of aromatic amino acids (via shikimate), which is also present in human intestinal bacteria, thus generating an additional form of damage to human health by affecting the intestinal microbiota, for which there is abundant evidence of our interdependence.^16^.

Due to the previous discussion, health professionals consider that there is an increase in risk and environmental diseases, which require confirmation by epidemiological studies: classic, descriptive and analytical, integrated into new scientific designs that cover systemic complexity.^17,18^. Designs that analyze health as an emergent state that arises from hierarchical network interactions between forces influencing the state of the system (dispersing, autopoietic or rigid), are needed^19,20^. These forces are expressed in historical, structural and contextual variables of health determinants and can be described in a systemic conceptual model to facilitate decisions taken by governments and people themselves^21^. To construct the model, available quali-quantitative data are systematized and missing data is covered by Participative-Action-Research (PAR) that includes all participants, from the problem formulation to interpreting results and discussing solutions^22^.

Since 2013, the Population Health Research Program (PISA) of the Hospital Italiano de Buenos Aires has investigated environmental risk factors, integrating basic, clinical-epidemiological and social science research for the design of public policies^23-26^. In June 2022, after validating the measurement of glyphosate in urine^26^, PISA was contacted by researchers from 9 de Julio (Buenos Aires) concerned over the environmental risks in their town. A health research scholarship from the Health Ministry made this study feasible.

This study aims to relate the presence of glyphosate in the human body with cultural, social and economical factors that influence rural locations in Argentina. The purpose of this research was to contribute scientific information to the French community and to the municipal council, to create health policies.

## Materials and methods

### Design

PAR^22^ with methodological triangulation^17^.

The study was carried out in a city named French of approximately 750 inhabitants (Latitude: -35.51; Longitude: -61) belonging to “9 de Julio” county in Buenos Aires district, Argentina.

People aged 18 years or older were included, who reported being local residents and having slept in the town. The initial sample size was 58 people, estimated on the basis of 20% frequency of glyphosate in urine, with a 95% confidence limit and 10% semi-amplitude. The sample was increased by 25% considering a response rate of 75%.

The probabilistic sampling was in 3 stages: (1) simple random sampling of blocks based on Google Maps; (2) systematic sampling of homes within the selected blocks; and (3) random selection among co-living family/group. Of a total of 46 blocks, 23 (50%) were selected. In each block the number of homes was registered, one of the four corners of the block was randomly chosen and the first home found, walking clockwise, was visited. A home member was randomly recruited using a dice. The procedure was repeated till the sample was completed^1^. Field work was completed in two days requiring 4 groups of people, each of which had 2 epidemiologists and a local citizen.

### Data collection

a 20 mL sample from morning’s first urine was requested. Samples were divided in two tubes (creatinine and glyphosate measurements), kept in dry ice until transported to the laboratory within 24 hs. Home, sociodemographic, environmental, occupation and health characteristics were evaluated with a validated questionnaire taken from the permanent home survey (INDEC, 2014); and environmental exposure questions came from environmental clinical history of the Health Ministry of the City of Buenos Aires. The variables were compounded in a RedCap form and were completed by telephone interviews. Environmental health perception was obtained from in-depth interviews of positive cases and from dialogue tools from social interaction.

### Analysis

glyphosate in urine was analyzed by the validated liquid chromatography linked to mass spectroscopy technique^26^, with results expressed in μmol/mol of urine creatinine with quantification values > 0.5 μg/L.

STATA_v13 was used for statistical analysis. Mean with standard deviation, median with 25-75 interquartile range, or in percentage and 95% confidence interval (95%CI), as necessary. Statistical significance was for P ≤ 0.05. Positive cases and environmental risk activities were geo-referenced.

Health determinant dimensions were analyzed with Atlas-ti v23 and were plotted based on Pérodeau’s model in discussion/consensus stages of the qualitative and quantitative collected data^21^.

Informed consent was obtained following the Province of Buenos Aires’ health regulation (law 11044). The protocol was approved by the Ethical Committee of Ministry of Health of Buenos Aires District (ACT-2022-32658194-GDEBA-CECMSALGP; PRIISA 8103) and the Helsinki declaration guidelines were followed.

## Results

### PAR process

There were three field trips:

1. March 2023: a face-to-face meeting was held with the Mayor and his team in *9 de Julio*. It was decided to carry out the research in the town of French. A mixed research team (MRT) was formed between local participants and researchers from the intervening institutions. Consensus was reached on the sampling framework, local communication and fieldwork planning^2^, following a Problem Based Participative Management (PBPM) method validated in hospital management^27^.
2. June 2023: samples were collected. In an open meeting held at a local community center, 50 people (citizens, *Guardianes de la Ecología, ConCiencia Agroecológica de 9 de Julio, Sociedad Italiana*, Elderly Care Home, Retirement Center and journalists) discussed different environmental risk sources in the town. Next, the Municipal Council made a formal request of the results and the media installed the topic as a social interest. Both instances drove the treatment of this topic on the political decision agenda.
3. December 2023: results were received by the Municipal Council and presented face-to-face to participants, thanks to previous communication and adequate preparation of the Retirement Center by the MRT.

### Socio-demographic profile

23 of the total 46 blocks were selected (50%), a percentage that was maintained in the “home” stratum: 76 homes from 23 blocks were selected; in total there were 152 homes in 46 blocks. All 76 chosen people provided a urine sample. Average age was 56.6 (16.2) years, 67% were women and 59 (77.6%) people completed the survey. The socio-demographic profile and risk factors are shown in Table 1; there were no significant differences between age and gender. Approximately half of them work and the rest are retired. In 21.8% of the homes, a history of oncological disease was reported, either the surveyed person or home dwellers. Of the surveyed people, 41.38% were covered by the national health-aid system for retirees and 28.8% perceive their health as regular or bad.

**Table 1:**
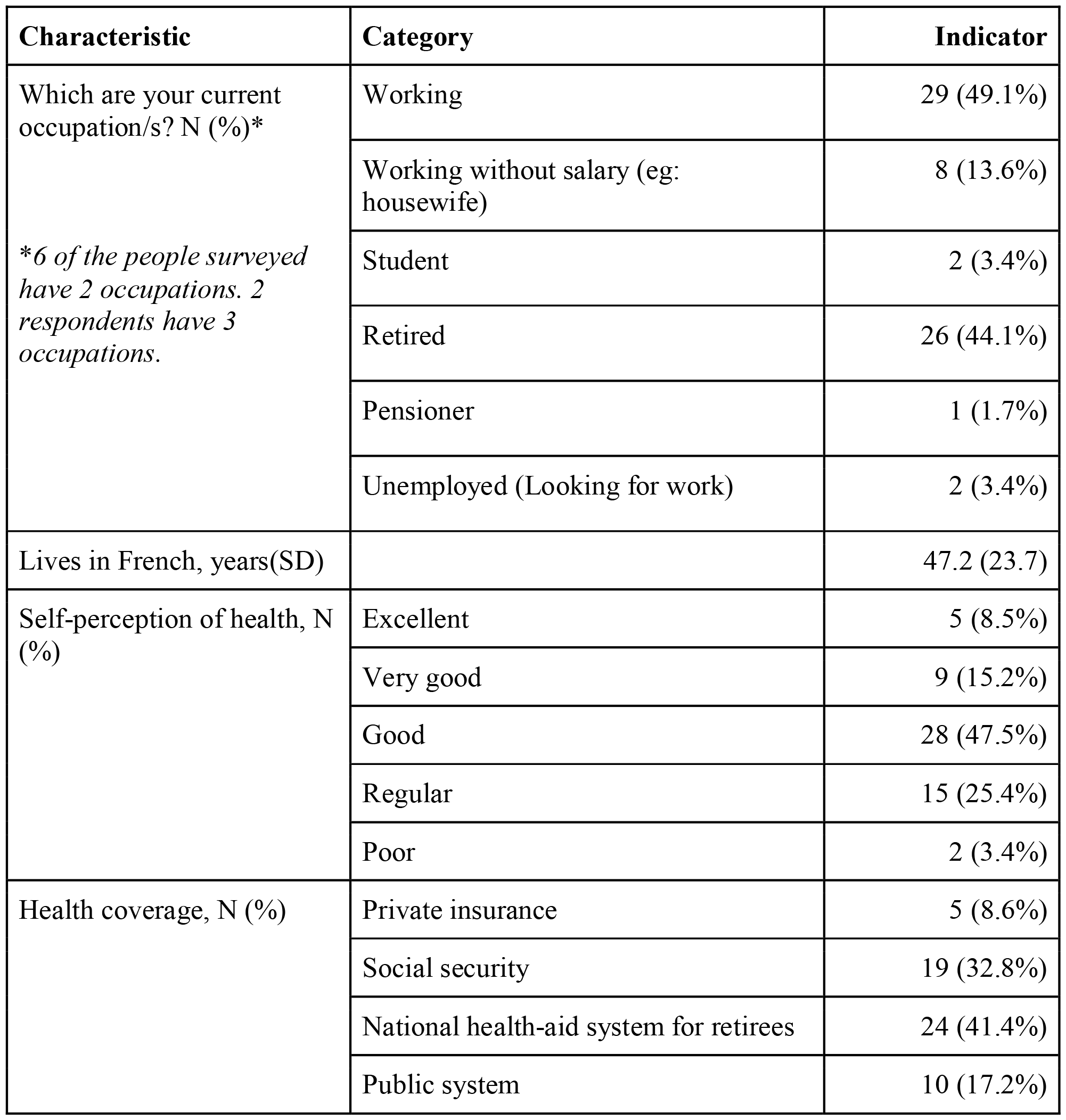
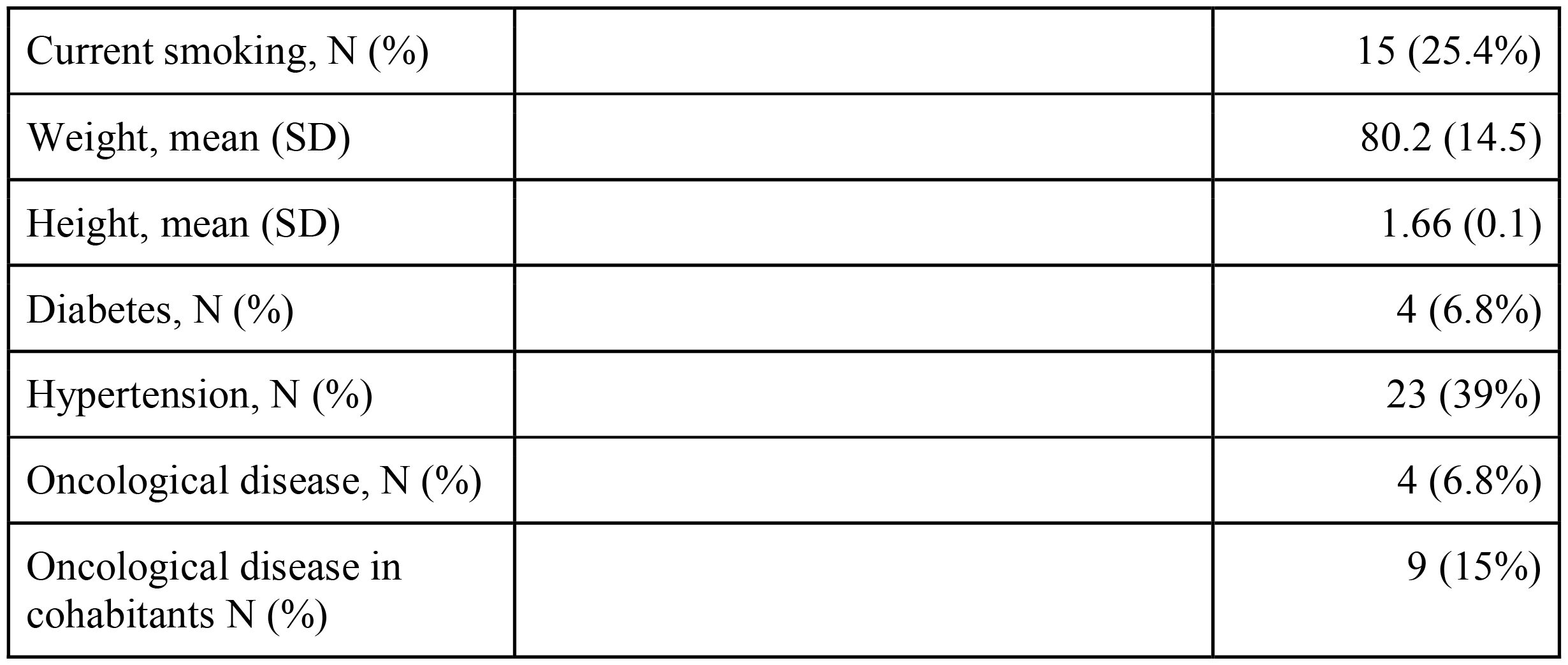
Sociodemographic profile and classic risk factors of the people surveyed in French. 2023.

### Urine glyphosate

13% (95%CI 6.5-23) of samples were quantifiable for glyphosate. The median concentration was 0.27 μmol/moles of creatinine; 25-75 interquartile range: 0.22-0.36; minimum of 0.18 and a maximum of 3.26 (Graphs 1). In all cases exposure related to work was discarded.

### Geo-referencing

Positive cases live close to the school: in the same block, on a side block and in a corner in front. The person with the highest level lives very close to the agrochemical warehouse of a grain company, having a concentration almost 3 standard deviations higher than the average.

### Self-perception of ways of contamination

The interviews revealed self-perception of ways of environmental contamination, identifying specific sources of exposure to agrochemicals within the urban radius: storage of seeds in silos, grain company and agrochemical warehouses.

> *“Crossing the railway line there is a warehouse where they have the chemicals. We had to ask that company to please close the gate and put something up because we can’t breathe even inside the room”*
>
> *“Here I have a grain company one block away… Each unloading of the truck is a whole blanket that releases that contaminated dust that has the cereal*”

There seems to be a certain resignation in having to put up with frequent odors of chemicals, dust and others related to the agricultural activity of the town.

> *“No, not every so often, it is felt often, early in the morning; if they fumigate, the wind brings, also at night time, or in the late afternoon. There is also a lot of pork smell, from the pigsty”*

The fumigation trucks circulate in the town and repair garages are also in the town.

> *“They pass there, all the fumigation trucks by the front door.”*

However, given that none of the people whose urine sample tested positive referred to have been specifically exposed to glyphosate or other pesticides, it is possible to argue that they are unaware of the specific circumstances of their own exposure, allowing the exposure to be considered environmental and not occupational, not being feasible to design preventive measures at the individual level of good practices for the use or care of agrochemicals.

Passive adaptation and helplessness to environmental exposure is observed:

> *“This summer, for example, it was so hot that in the evening you opened the windows. And I don’t know the chemicals or anything, but the smell was so strong. It came into the house and you had to close the windows again.”*
>
> *“There are clandestine sprayings by night, and if you call the police they don’t come”. “If we carry on like this the earth will be sand dunes.”*

### Systemic determinants of health

The systemic model (Figure 1) showed structural and contextual variables that account for the expression of a historical evolution of development forces, with few expressions of care forces:

**Figure 1:**
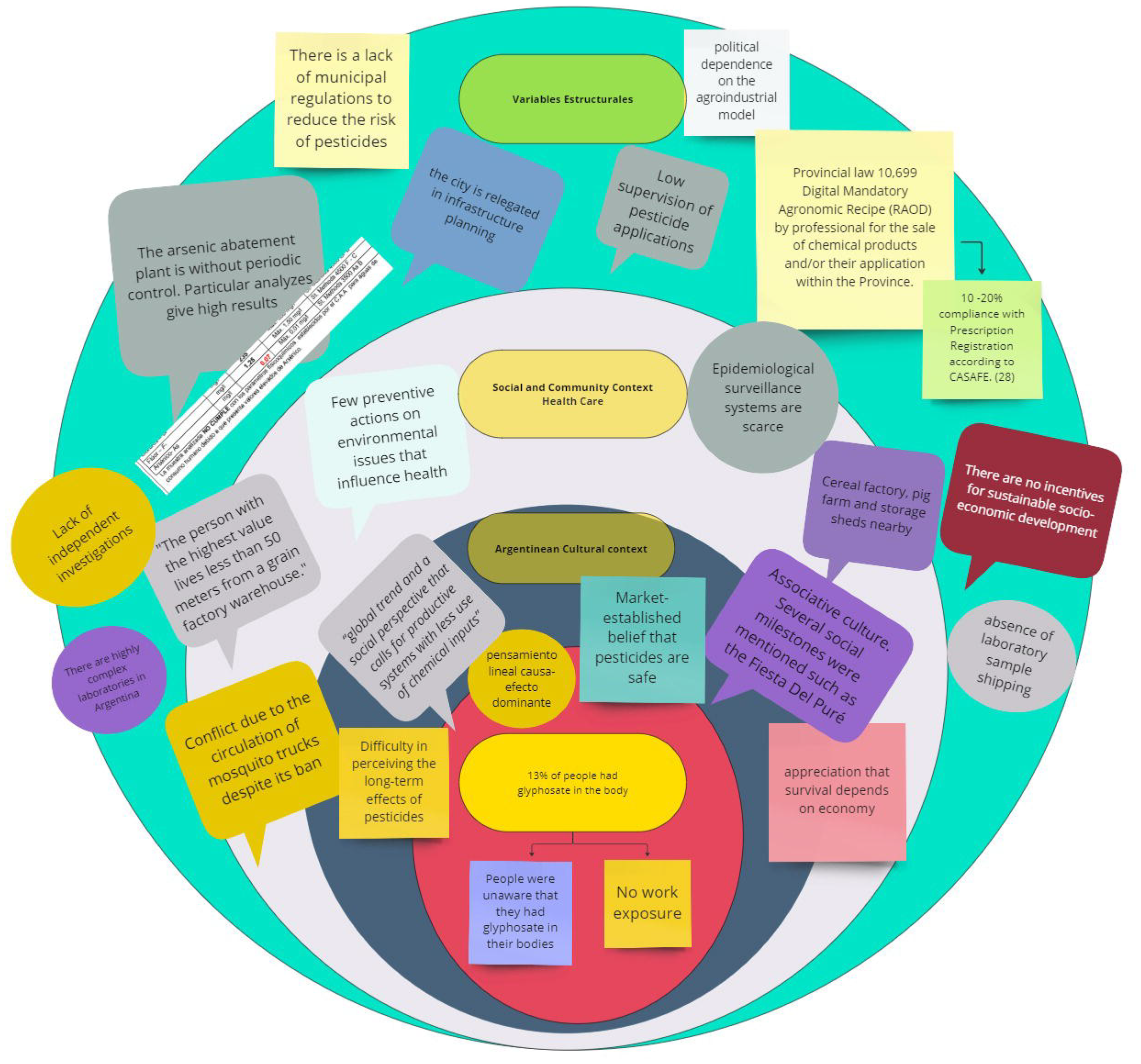
Systemic model of structural, contextual and cultural conditions that directly or indirectly influence people’s environmental pollution in French.

At the structural level:

a. The arsenic abatement plant does not have periodic controls. According to the UTN-FTR laboratory conducted by a private individual in 2021, it is not fit for consumption (exceeds 7 times the accepted limit).
b. It is observed and reported that the town is neglected in infrastructure planning. The town has no public transportation. The train station is not active despite complaints.
c. The sanitary infrastructure, and the health center, do not cover the needs of a long-lived population. No preventive actions were mentioned by the health system on the socio-environmental problems affecting health.

At the contextual level:

a. The insufficient regulation of the production model is reflected in the current laws due to their lack of effective implementation. An example of this is the Provincial Law 10,669, that prohibits the application of products without a mandatory agronomic prescription. According to the Agricultural Sanitation and Fertilizer Chamber (CASAFE, acronym in Spanish). This regulation is fulfilled by 20% ^28^.
b. The “culture of scarce regulation and of evading control” is evidenced by the presence of pesticide storage warehouses, a grain plant, pig farms without wastewater treatment, and an open dump, all within the limits of the town.
c. Environmental health care is promoted, partly by the local school, by young people and by new inhabitants that promote culinary developments in rural tourism business^2^.

## Discussion

The population design of the study allows us to infer that the implications of agricultural activity on the human body are manifested in 13% of the French population due to the presence of glyphosate in the urine. Of the approximately 750 inhabitants that live in the town, approximately 100 would have glyphosate due to environmental exposure during similar days of our study. We believe that having a probabilistically representative sample is a strength of the study and allows the results to be generalized to similar populations. Thus, a person living in a similar rural locality (9 de Julio and its six neighboring cantons have 43 rural localities with approximately 45 thousand inhabitants in total), would have a 13% probability of having glyphosate in urine.

The qualitative results support that the data obtained in June (considered low season for pesticide spraying) were due to environmental and non-occupational exposure. Glyphosate is eliminated from the body between 48-72 hours. This means that if sampling had been repeated on the same population a few days later, and the exposure conditions were maintained, a person who was positive in a first sampling could be negative in a subsequent one and vice versa; with a 95% chance of percent positives being between 6.5 and 23%. We consider that an additional study should be carried out during the peak fumigation season (October or November) since environmental and working conditions could differ.

It is important to note that in the current scientific debate, focusing on the levels found in urine is questioned, since there would be no “accepted normal level” in light of the evidence of transgenerational epigenetic and endocrine effects in laboratory animals.^6^. This evidence cannot be ethically documented in humans, as it would require a long longitudinal study including a similar unexposed group. Being an environmental factor, it means that the entire population is at risk of exposure in some way, which means that it is possible to have an unexposed group.

Although the reductionist concept of risk based on dose-response curves is questioned, we share that in 8 studies the values of occupational exposure to glyphosate in urine were between 1.54 and 435 nmol/L; and in 14 studies carried out on different general population groups the values were between 1.54 and 45 nmol/L^6^. Our results are expressed as μmol/mole of creatinine (relative to a renal variable); but for comparative reasons, the mean value expressed in nmol/L not normalizing by creatinine value is 5.7 (SD 5.3), which is within the range of other studies.

Georeferencing showed that the positive cases were close to the school; Given that children are more vulnerable to epigenetic effects and endocrine alterations, it is suggested to measure a sample of schoolchildren. 22% of homes reported a co-habitant having an oncological illness in a population whose average age was 60 years, which indicates high care and attentions requirements (diagnosis, treatment, contention care, etc.).

Although there are highly complex laboratories that measure pesticides, referral circuits for acute poisonings and/or exposure monitoring, whether environmental or occupational, do not have a protocol in the Argentinean health system. To begin, we propose to computerize community surveillance systems (as is already done with application notifications), for suspected exposure and for possible diseases related to pesticides (through a cell phone app available to people). This initiative requires financing and we consider that must be of interest to the global sustainability agenda. The appropriation by the community of the research results for the design of a website (https://frenchsaludambiental.com.ar/) for future actions was thanks to participatory tools under the PAR design.

From systems theory, the presence of a substance in urine, which is only used for agricultural activity in the town, indicates that a higher level of organization of the system (economic interactions) influences a lower evolutionary level (biological interactions). For the system to evolve without detriment to its parts and/or levels, the forces must be in balance^18,19^. In the systemic model of Figure 1, we observe that the interrelation between variables at different levels allows us to consider that the forces of dissipation of the system —those that in the long term affect sustainability— are influenced by the productive model without being antagonized by forces of care —by regulations, whether of cultural community or state origin—^18^. According to Teeple (2000), the evolution of “forces which only generate capital”, has led to an unequal and dystopic system development because some parts (corporations) grew entropically using the energy of the other parts of the system. The reproduction of these forces in human interaction would lead to greater global social inequality and environmental damage^29^. Systems care actions, under One Health Approach, could lead to an equilibrium. Combining the points of view and collaboration between scientific institutions, government organizations and the civil society institutions is the challenge to promote sustainable changes in French’s health. Institutionalized democratic processes for health sustainability were promised by the deliberative council of 9 de Julio after these results were obtained.

Summing up, in the systemic approach, the butterfly effect has a mathematical demonstration on how small changes in a part of the system can generate global changes on the whole^30^. We consider that this research is a contribution in the salutogenic direction, reflecting the generative power of biocentric forces. Quoting a local citizen: *“We are surprised that we can achieve something like this in our city. I’m surprised anyone cares about us. It also moves me that this little bit could be the beginning of something more*.”

## Conclusion

During the low fumigation season, in rural populations of Buenos Aires, Argentina, there are people who have glyphosate in their urine. This finding is a consequence of the effect of agricultural activities on the human body, responding to systemic forces that lead to the passive, involuntary and chronic absorption of environmental chemicals by humans. The pressure to continue growing economically is not balanced by individual, social or state forces of care. As the effect of environmental exposure is global, individual care measures to avoid the presence of glyphosate in urine are not enough, unless the person migrates from the town.

## Data Availability

All data produced in the present study are available upon reasonable request to the authors

https://frenchsaludambiental.com.ar/

---

**Graph 1:**
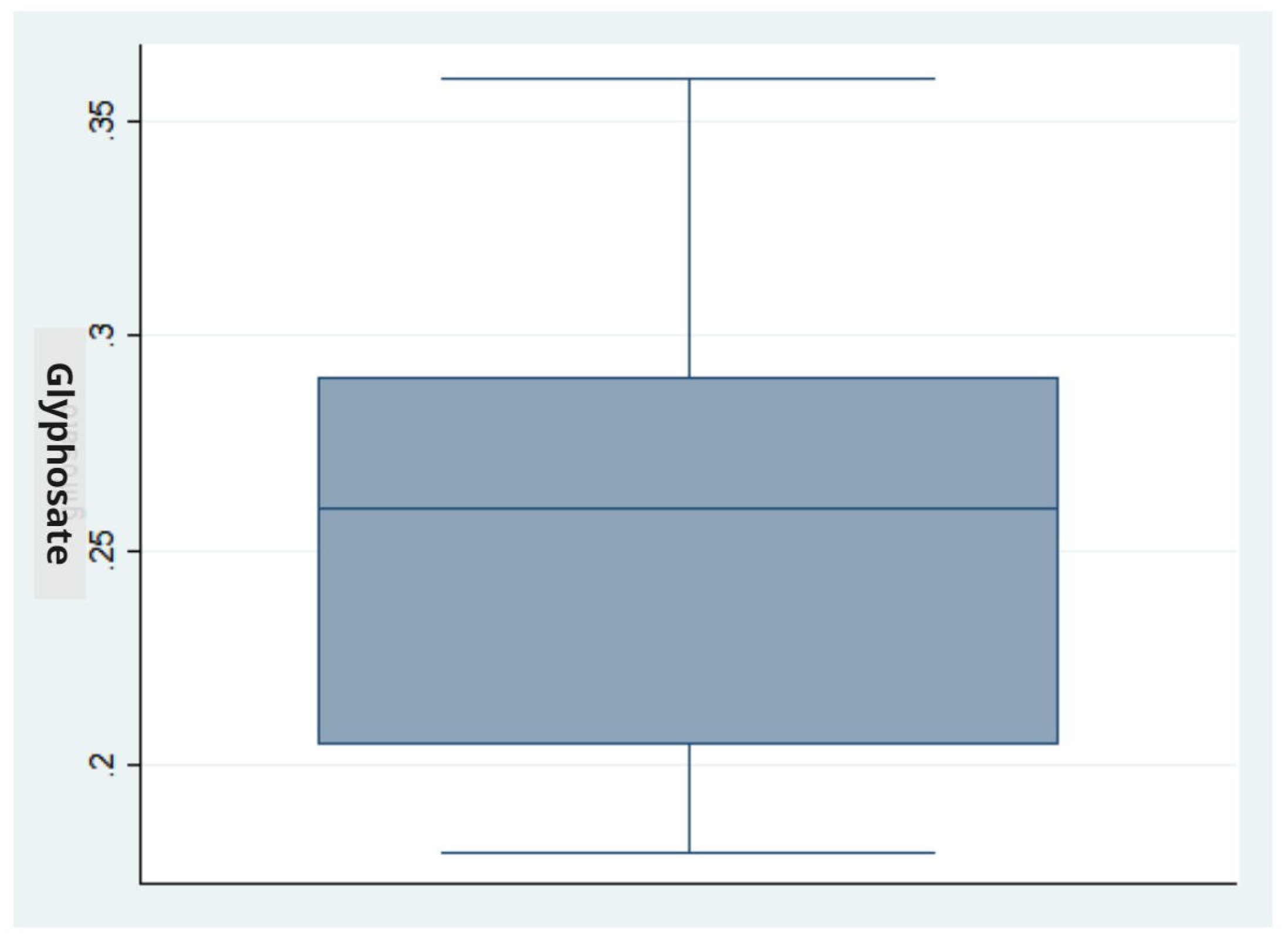
Box Plot of urine glyphosate values in quantifiable samples (n 8, without outliers). French. Year 2023.

## Footnotes

1 Annex I: complementary information can be provided by the author on request

2 https://www.lanacion.com.ar/sociedad/nos-animamos-a-cumplir-un-sueno-vendieron-todo-y-dejaron-la-ciudad-para-reabrir-un-historico-nid13082021/

